# The characteristics and death risk factors of 132 COVID-19 pneumonia patients with comorbidities: a retrospective single center analysis in Wuhan, China

**DOI:** 10.1101/2020.05.07.20092882

**Authors:** Chen Chen, Jingyi Zhang, Chang Li, Zhishuo Hu, Ming Zhang, Pei Tu, Lei Liu, Wenxia Zong

## Abstract

**Background:** The new coronavirus pneumonia (COVID-19) has evolved into a global pandemic disease, and the epidemiological characteristics of the disease have been reported in detail. However, many patients with new coronary pneumonia have comorbidities, and there are few researches reported in this special population.

**Methods:** a retrospective analysis was performed on 132 consecutive COVID-19 patients with comorbidities from January 19, 2020 to March 7, 2020 in Hubei Third People’s Hospital. Patients were divided into mild group and critical group and were followed up to the clinical endpoint. The observation biomarkers include the clinical feature, blood routine, blood biochemistry, inflammation biomarkers, and coagulation function. Univariate and multivariate logistic regression was used to analyze the risk factors associated with death.

**Results:** 132 patients were enrolled in this study and divided into the mild group (n=109, 82.6%) and critical group (n=23, 17.4%), of whom 119 were discharged and 13 were died in hospital. The all-cause mortality rate was 9.8%, of which 7 patients died of respiratory failure, 5 patients died of heart failure, and 1patient died of chronic renal failure. There was significant statistical difference of mortality rates between the mild group (5.5%) and the critical group (30.4%). The average time of hospitalization was 16.9 (9, 22) days. Hypertension was the most common comorbidity (n=90, 68.2%), followed by diabetes (n=45, 34.1%), coronary heart disease (31, 23.5%). Compared with the mild group, the patients were older in critical group (P <0.05), and neutrophils, neutrophil ratio, neutrophil-lymphocyte ratio (NLR), serum urea nitrogen (BUN), procalcitonin (PCT), C-reactive protein CRP), serum amyloid protein (SSA), N-terminal brain natriuretic peptide precursor (NT-pro BNP) were significantly increased (P <0.05). However, lymphocytes lymphocyte ratio, albumin were lower than those in the critical group (P <0.05). The patients were further divided into the survivor group (n=119, 90.2%) and the non-survivor group (n=13, 9.8%). Compared with the survivor group, the death rate of patients with coronary heart disease was significantly increased (53.8% vs 20.2%), and The neutrophil ratio, aspartate aminotransferase (AST), BUN, PCT, CRP, SAA, interleukin-6(IL-6) and D-dimer were significantly increased (P <0.05), while the lymphocytes and NLR reduced (P <0.05). Multivariate logistic stepwise regression analysis showed that the past medical history of coronary heart disease[OR:2.806 95%CI:0.971~16.795], decreased lymphocytes [OR: 0.040, 95%CI:0.001~2.306], increased AST[OR:1.026, 95%CI:1.000~1.052], increased SSA[OR:1.021, 95%CI:1.001~1.025], and increased D-dimer[OR:1.231, 95%CI:1.042~1.456] are risk factors associated with death in COVID-19 patients pneumonia with comorbidities.

**Conclusion:** The mortality rate of COVID-19 patients with coronary heart disease is relatively high. In all patients, the lower lymphocytes, and higher NLR, BUN, PCT, CRP, SSA, D-dimer are significant characteristics. The past medical history of coronary heart disease, decreased lymphocytes, increased AST, SSA and D-dimer are risk factors associated with death in COVID-19 patients’ pneumonia with comorbidities

## Introduction

Since the outbreak of new coronavirus pneumonia COVID-19, more than 800,000 people have been infected worldwide, far larger than the SARS in 2003, and it has evolved into a global pandemic(*1*). Such diseases are mainly transmitted through respiratory droplets and close contact, and the clinical features of different individuals vary greatly (*2, 3*). At the same time, there are data showing that about one-quarter of patients with new coronary pneumonia in China currently have comorbidities. The presence of comorbidities can easily lead to a poor clinical prognosis, contributing to rapid progression of the disease to acute respiratory distress syndrome, respiratory failure and even death. Compared with patients without comorbidities, as long as a patient has a comorbidity, the risk of poor prognosis will increase by about 79% (*4*). Previous studies have shown that among patients with H7N9 infection, any comorbidities may lead to a 3.4-fold increased risk developing into acute respiratory distress syndrome (*5*). Therefore, in the case of pneumonia patients with other diseases, the treatment of comorbidities is also very important. This article summarizes the characteristics of 132 cases of patients diagnosed with COVID-19 combined with other comorbidities, in order to provide some ideas for the prevention and treatment of patients with comorbidities.

## Methods

### Study population

459 consecutive cases were from Hubei Third People’s Hospital from January 19, 2020 to March 7, 2020, of which 132 patients with comorbidities were enrolled for this study (accounting for 28.8%). There is a retrospective study of 132 patients, all enrolled patients were diagnosed with COVID-19 pneumonia according to WHO interim guidance (*6*). 132 patients were divided into two groups: the mild group and the critical group. This study was approved by the Ethics Committee of Hubei Third People’s Hospital and the hospital granted a waiver of informed consent from study participants.

### Procedure

To identify COVID-19 infection, throat swab samples were obtained from all patients at admission and tested using real-time reverse transcriptase–polymerase chain reaction assays according to the same protocol described previously (*7*). All patient’s data was stored in a locked, password protected computer, and the majority of the clinical data used in this study was collected from the first day of hospital admission indicated otherwise. Common observation biomarkers including blood routine, lipid profiles, liver damage indices, renal dysfunction indices, coagulation function indices, and inflammatory biomarkers and other biomarkers were measured by standard laboratory techniques.

### Statistical analysis

Statistical analyses were performed using SPSS 22.0 for Windows (SPSS, Chicago, Illinois). Continuous variables were expressed as mean (standard deviation, SD) in the case of normal distribution, and differences were determined by Student t test. Data were expressed as a median (25th and 75th percentiles: P25, P75) in the case of skewed distribution and were compared using the Mann-Whitney U test. Categorical variables were presented as a number and percentage and were compared using the χ^2^ test. To explore the risk factors associated with in-hospital death, multivariate logistic regression models were used. Considering the total number of deaths (n=13) in our study and to avoid overfitting in the model, the variables that were considered significant in previous univariate analysis were chosen for multivariate analysis. The results of logistic regression analysis were presented as an odds ratio (OR) and 95% confidence interval (CI). The A 2-tailed *P* < .05 was considered significant.

## Result

### 1. Demographics and Clinical feature

132 patients diagnosed with COVID-19 pneumonia with comorbidities were enrolled, accounting for 28.8% of the patients diagnosed with COVID-19 pneumonia (n=459) in our hospital during the same period, including 109 cases in the mild group and 23 cases in critical group. The age of enrolled patients with comorbidities was 63.4 years (56, 71). 76 were males (57.6%) and 56 were females (42.4%) respectively. The most common symptom was fever(65.9%), the same as cough or sputum(65.9%). Followed by the fever was fatigue with anorexia(34.8%). and both 19.7% of patients had dyspnea and palpitations with chest tightness. The average time of hospitalization was 16.9 (9, 22) days, and there was no significant difference between the mild group and the critical group (P = 0.209). Hypertension was the most common comorbidity (68.2%), followed by diabetes (34.1%), coronary heart disease (23.5%), and hyperlipidemia (14.4%). Patients with stroke and chronic kidney disease only were 5.3% and 4.5% respectively. The highest proportion of drug used for treatment during hospitalization was antibiotics (77.3%), followed by Arbidol hydrochloride (60.1%) and 40mg methylprednisolone twice a day(49.2%). The percentage of using Oseltamivir and Ribavirin was 34.1% and 27.3%. The all-cause mortality rate was 9.8%, of which 7 patients died of respiratory failure, 5 patients died of heart failure, and 1patient died of chronic renal failure. There was significant statistical difference of mortality rates between the mild group (5.5%) and the critical group (30.4%) (P <0.05).

### 2. Laboratory Parameters between the mild group and the critical group

Laboratory findings between the mild group and the critical group on hospital admission are summarized in Table 2. Compared with the mild group, the critical group demonstrated increased neutrophil ratio and NLR, while decreased lymphocyte ratio and lymphocytes. Patients presented with lower albumin and elevated BUN, BNP, PCT, CRP, SAA and FDP in the critical group. While platelet count, hemoglobin, myocardial enzyme profile and lipid metabolism indices were similar between the two groups.

**Table 1.** Demographics and Clinical feature of COVID-19 patients with commodities on admission

|  | Total<br>(n=132) | Mild cases<br>(n=109) | Critical cases<br>(n=23) | *P* value |
| --- | --- | --- | --- | --- |
| Characteristics |  |  |  |  |
| Age, years | 63.4(56-71) | 62.4(52.5-70) | 68.3(63-79 ) | 0.025 |
| Sex |  |  |  | 0.152 |
| Male | 76(57.6%) | 61(56.0%) | 15(65.2%) |  |
| Female | 56(42.4%) | 48(44.0%) | 8(34.8%) |  |
| Duration in hospital,<br>days | 16.9(9-22) | 15.9(7-20) | 17.1(10-22) | 0.209 |
| Initial symptom |  |  |  |  |
| Fever<br>(temperature≥37.3℃) | 87(65.9%) | 69(63.3%) | 18(78.2%) | 0.169 |
| Cough or sputum | 87(65.9%) | 69(63.3%) | 23(100%) | 0.087 |
| Dyspnea | 26(19.7%) | 20(18.3%) | 6(26.6%) | 0.668 |
| Palpitation | 25(18.9%) | 21(19.2%) | 4(17.2%) | 0.132 |
| Fatigue or anorexia | 46(34.8%) | 40(36.7%) | 6(26.1%) | 0.079 |
| Diarrhea | 17(12.9%) | 14(12.8%) | 3(13.0%) | 0.921 |
| Others | 6(4.5%) | 5(4.6%) | 1(4.3%) | 0.865 |
| Comorbidity |  |  |  |  |
| Hypertension | 90(68.2%) | 71(65.1%) | 19(82.6%) | 0.102 |
| Coronary heart<br>disease | 31(23.5%) | 24(22.0%) | 7(30.4%) | 0.387 |
| Hyperlipidemia | 19(14.4%) | 16(12.1%) | 3(13.0%) | 0.839 |
| Diabetes | 45(34.1%) | 36(27.3%) | 9(39.1) | 0.575 |
| Stroke | 7(5.3%) | 6(4.5%) | 1(4.3%) | 0.822 |
| Chronic kidney<br>disease | 6(4.5%) | 6(4.5%) | 0(0) | 0.249 |
| Other | 5(3.8%) | 5(3.8%) | 0(0) | 0.295 |
| Drug usage |  |  |  |  |
| Arbidol<br>hydrochloride | 80(60.1%) | 67(61.5%) | 13(56.5%) | 0.659 |
| Oseltamivir | 45(34.1%) | 34(31.1%) | 11(47.8%) | 0.126 |
| Ribavirin | 36(27.3%) | 28(25.7%) | 8(34.8%) | 0.374 |
| Methylprednisolone | 65(49.2%) | 44(40.4%) | 21(91.3%) | < 0.001 |
| Antibiotics | 102(77.3%) | 80(73.4%) | 22(95.6%) | 0.021 |
| Gamma globulin | 42(31.2%) | 24(22.0%) | 18(78.3%) | < 0.001 |
| Albumin | 17(12.9%) | 10(9.2% ) | 7(30.4%) | 0.006 |
| Deaths | 13(9.8%) | 6(5.5%) | 7(30/4%) | < 0.001 |
| Respiratory failure | 7 | 3 | 4 |  |
| Heart failure | 5 | 2 | 3 |  |
| Renal failure | 1 | 1 | 0 |  |

**Table 2.** Laboratory findings of COVID-19 patients with comorbidities on admission

|  | Mild cases | Critical cases | P value |
| --- | --- | --- | --- |
| Blood routine |  |  |  |
| White cell count,<br>×10<sup>9</sup>/L | 6.40(4.22-7.85) | 5.83(3.95-8.94) | 0.888 |
| Red blood cell count<br>10<sup>12</sup>/L | 4.10(3.76-4.45) | 4.19(3.66-4.49) | 0.867 |
| Hemoglobin, g/L | 128(116-139) | 127.50(107.00-140.75) | 0.824 |
| Platelet count ×10<sup>9</sup>/L | 225(158.75-279.75) | 171.50(107.00-273.25) | 0.058 |
| Neutrophil ratio (%) | 73.95(61.85-83.90) | 84.00(70.59-90.33) | 0.008 |
| Neutrophil count<br>×10<sup>9</sup>/L | 4.54(2.82-5.96) | 5.10(2.86-7.67) | 0.481 |
| Lymphocyte ratio (%) | 18.45(11.22-27.28) | 11.85(6.75-20.10) | 0.006 |
| Lymphocyte count<br>×10<sup>9</sup>/L | 0.95(0.70-1.47) | 0.69(0.56-0.97) | 0.003 |
| Neutrophil-lymphocyte<br>ratio (NLR) | 4.10(2.19-7.51) | 7.08(3.48-12.89) | 0.010 |
| Eosinophil ratio (%) | 0.50(0.10-1.70) | 0.10(0-0.80) | 0.096 |
| Eosinophil count<br>×10<sup>9</sup>/L | 0.03(0-0.10) | 0.01(0-0.08) | 0.226 |
| Monocyte ratio (%) | 7(4.50-8.63) | 4.85(3.10-8.83) | 0.054 |
| Monocyte count<br>×10<sup>9</sup>/L | 0.41(0.28-0.55) | 0.33(0.20-0.44) | 0.055 |
| Blood biochemical index |  |  |  |
| ALT,U/L | 26(18.78-40.13) | 23.30(16.45-54.90) | 0.776 |
| AST,U/L | 36.25(27.55-45.25) | 39.80(27.00-95.15) | 0.776 |
| Albumin, g/L | 37.35(33.73-42.50) | 33.70(31.70-35.60) | 0.001 |
| BUN, mmol/L | 4.48(3.44-5.89) | 5.80(4.54-7.710) | 0.006 |
| Creatinine, umol/l | 61.50(52.05-76.01) | 71.00(49.50-129.50) | 0.149 |
| Uric acid, umol/l | 280.50(210.00-377.00) | 248(217.50-404.00) | 0.760 |
| CK,U/L | 69(46.10-133.00) | 75.50(36.25-501.75) | 0.296 |
| CK-MB,U/L | 11.10(8.20-16.56) | 8.10(5.37-56.48) | 0.143 |
| BNP, ng/L | 199(42-545) | 684.50(454.00-3659.25) | 0.017 |
| Inflammatory biomarker |  |  |  |
| PCT, ng/ml | 0.04(0-0.09) | 0.11(0-6.23) | < 0.001 |
| CRP, mg/L | 8.74(2.13-51.78) | 23.42(4.88-117.468) | < 0.001 |
| ESR, mm/h | 52.00(20.80-78.90) | 73.50(30.75-110.00) | 0.052 |
| SAA, mg/L | 59.95(6.29-463.18) | 421.75(34.88-939.68) | 0.002 |
| IL-6, pg/ml | 11.28(1.50-28.30) | 5.15(1.95-21.07) | 0.825 |
| K<sup>+</sup> | 3.79(3.59-4.28) | 3.9(3.22-4.62) | 0.615 |
| Blood lipid |  |  |  |
| TC, mmol/L | 4.53(3.46-5.26) | 4.40(2.93-4.95) | 0.108 |
| TG, mmol/L | 1.36(1.00-1.84) | 1.36(1.13-1.66) | 0.300 |
| LDL-C, mmol/L | 2.97(2.28-3.42) | 2.45(1.94-3.53) | 0.123 |
| D-dimmer, ug/ml | 0.69(0.33-1.35) | 2.30(0.60-9.97) | 0.064 |
| FDP, ug/ml | 4.47(2.72-5.54) | 6.09(4.78-13.94) | 0.011 |

### 3. Clinical Characteristics and laboratory results of COVID-19 patients between survivors and non-survivors

There was significant statistical difference of mortality rates between the mild group (5.5%) and the critical group (30.4%). Male patients accounted for 69.2% of deaths, and most of them were over 65 years old (69.2%). Among them, the highest proportion of combined with hypertension was 76.9%, followed by coronary heart disease (53.8%). Compared with the survivor group, the proportion of death cases with coronary heart disease was significantly higher in the non-survivor group, with a statistically significant difference (*P* <0.05), the same as the neutrophils in the dead group was significantly increased (*P* <0.05). At the same time, we observed that compared with survivor group, the patients in the non-survivor group demonstrated increased lymphocyte ratio, and NLR, while lower lymphocytes, eosinophils, monocyte ratio, and monocytes. AST and BUN of the patients in the non-survivor group were similarly significantly higher than those in the survivor group. Then we compared the differences of inflammatory factors between the two groups. Compared with the survivor group, the levels of PCT, CRP, SAA and IL-6 in the non-survivor group were significantly increased (*P* <0.01). In the coagulation indices, the patients in the non-survivor group showed increased D-dimer and FDP than those in the survivor group (*P* <0.05).

### 4. Univariate and Multivariate logistic Regression of Factors Associated with Progression to Death (Table 4)

Table 4 displays the results of univariate and multivariate logistic regression analyses. Covariates for multivariate analysis were selected based on clinical importance and predictors associated with each Non-survivor (P < 0.05) in univariate analysis. In multivariate logistic regression analysis, we found that the past medical history of coronary heart disease[OR:2.806 95%CI:0.971~16.795], decreased lymphocytes [OR:0.040, 95%CI:0.001~2.306], increased AST[OR:1.026, 95%CI:1.000~1.052], increased SSA[OR:1.021, 95%CI:1.001~1.025], and increased D-dimer[OR:1.231, 95%CI:1.042~1.456] are risk factors associated with death in COVID-19 patients pneumonia with comorbidities.

**Table 3.**
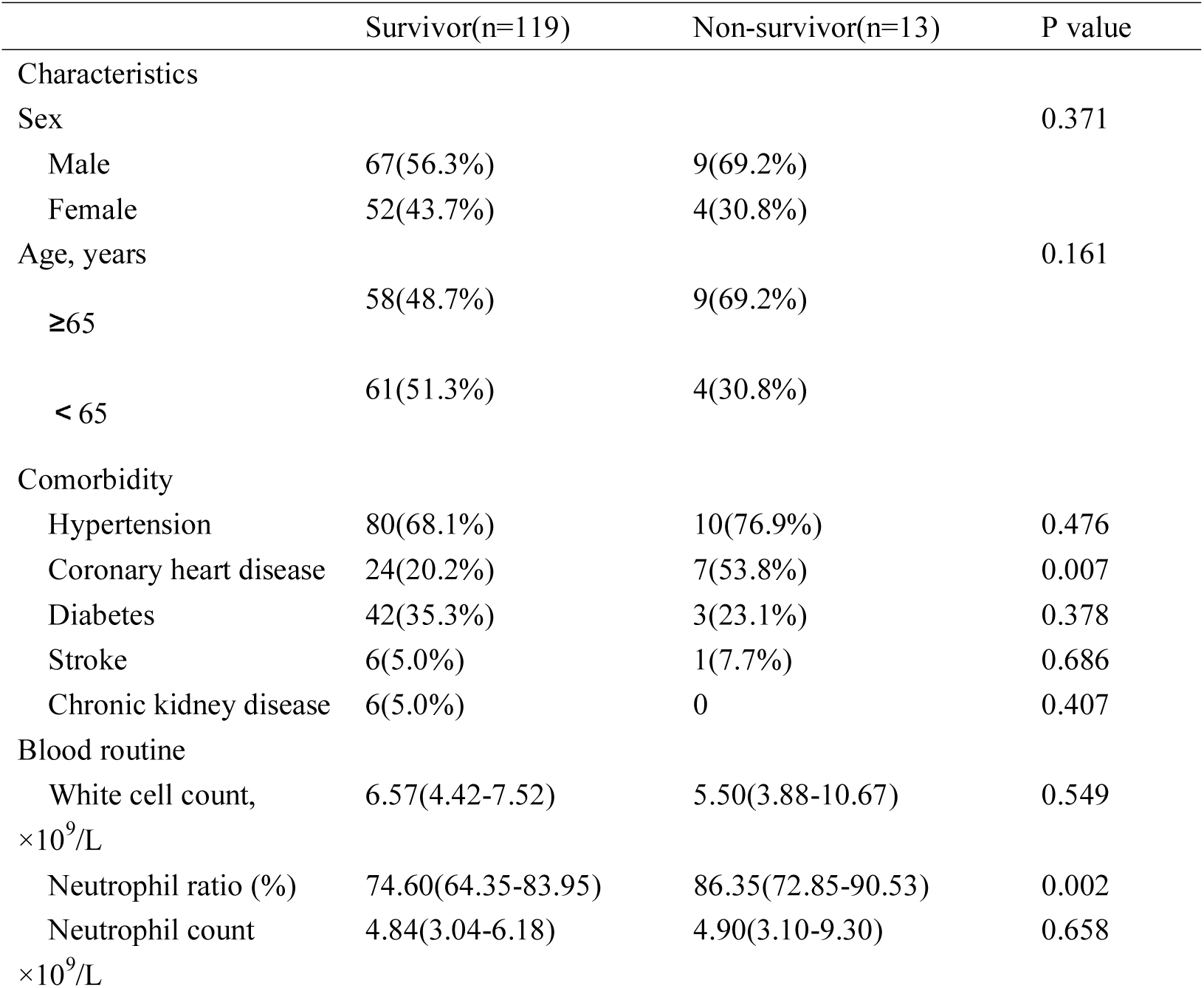
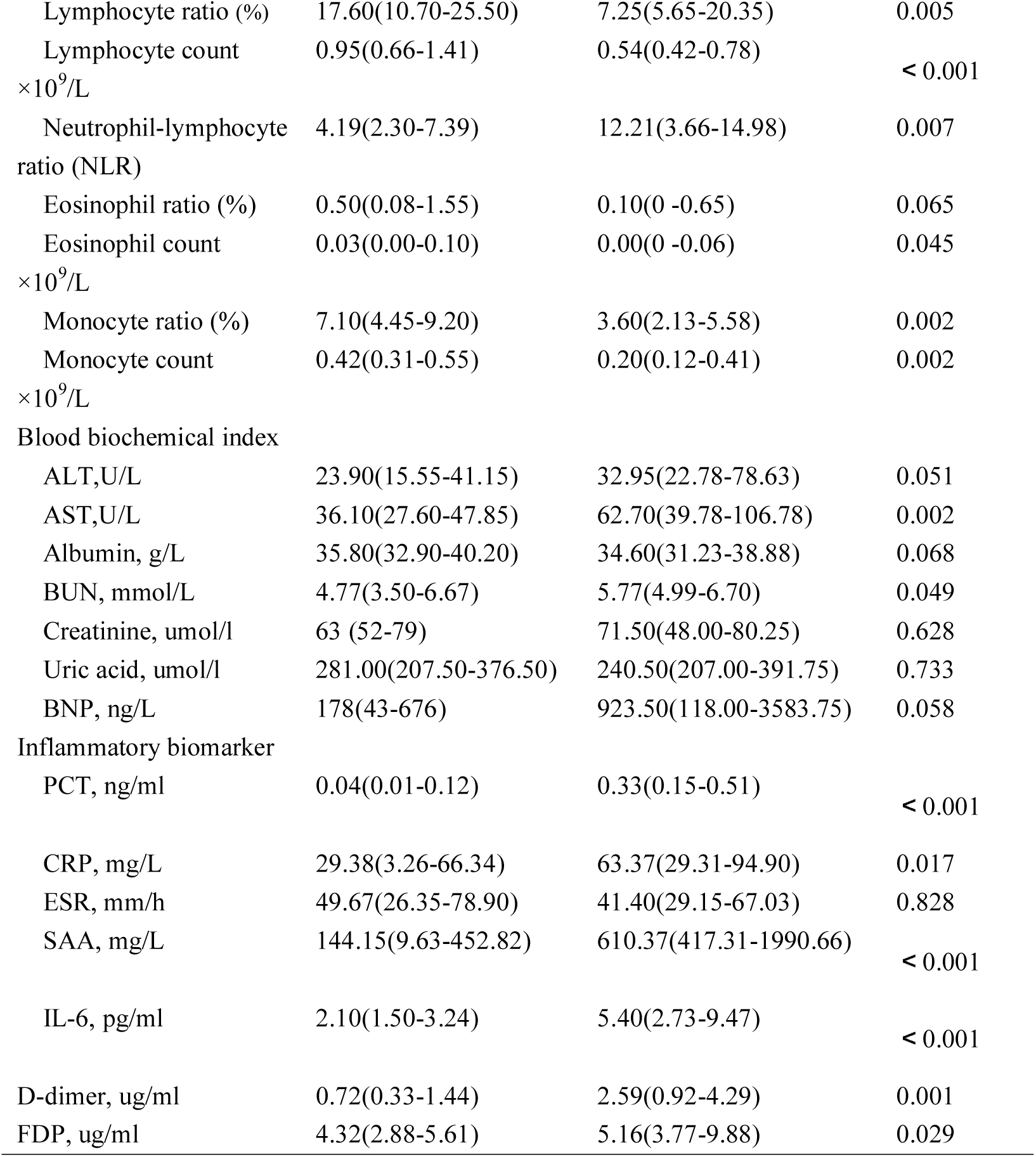
Clinical, laboratory findings of non-survivor and survivor patients with comorbidities on admission

|  | Survivor(n=119) | Non-survivor(n=13) | P value |
| --- | --- | --- | --- |
| Characteristics |  |  |  |
| Sex |  |  | 0.371 |
| Male | 67(56.3%) | 9(69.2%) |  |
| Female | 52(43.7%) | 4(30.8%) |  |
| Age, years |  |  | 0.161 |
| ≥65 | 58(48.7%) | 9(69.2%) |  |
| < 65 | 61(51.3%) | 4(30.8%) |  |
| Comorbidity |  |  |  |
| Hypertension | 80(68.1%) | 10(76.9%) | 0.476 |
| Coronary heart disease | 24(20.2%) | 7(53.8%) | 0.007 |
| Diabetes | 42(35.3%) | 3(23.1%) | 0.378 |
| Stroke | 6(5.0%) | 1(7.7%) | 0.686 |
| Chronic kidney disease | 6(5.0%) | 0 | 0.407 |
| Blood routine |  |  |  |
| White cell count,<br>×10<sup>9</sup>/L | 6.57(4.42-7.52) | 5.50(3.88-10.67) | 0.549 |
| Neutrophil ratio (%) | 74.60(64.35-83.95) | 86.35(72.85-90.53) | 0.002 |
| Neutrophil count<br>×10<sup>9</sup>/L | 4.84(3.04-6.18) | 4.90(3.10-9.30) | 0.658 |
| Lymphocyte ratio (%) | 17.60(10.70-25.50) | 7.25(5.65-20.35) | 0.005 |
| Lymphocyte count<br>×10<sup>9</sup>/L | 0.95(0.66-1.41) | 0.54(0.42-0.78) | < 0.001 |
| Neutrophil-lymphocyte<br>ratio (NLR) | 4.19(2.30-7.39) | 12.21(3.66-14.98) | 0.007 |
| Eosinophil ratio (%) | 0.50(0.08-1.55) | 0.10(0 -0.65) | 0.065 |
| Eosinophil count<br>×10<sup>9</sup>/L | 0.03(0.00-0.10) | 0.00(0 -0.06) | 0.045 |
| Monocyte ratio (%) | 7.10(4.45-9.20) | 3.60(2.13-5.58) | 0.002 |
| Monocyte count<br>×10<sup>9</sup>/L | 0.42(0.31-0.55) | 0.20(0.12-0.41) | 0.002 |
| Blood biochemical index |  |  |  |
| ALT,U/L | 23.90(15.55-41.15) | 32.95(22.78-78.63) | 0.051 |
| AST,U/L | 36.10(27.60-47.85) | 62.70(39.78-106.78) | 0.002 |
| Albumin, g/L | 35.80(32.90-40.20) | 34.60(31.23-38.88) | 0.068 |
| BUN, mmol/L | 4.77(3.50-6.67) | 5.77(4.99-6.70) | 0.049 |
| Creatinine, umol/l | 63 (52-79) | 71.50(48.00-80.25) | 0.628 |
| Uric acid, umol/l | 281.00(207.50-376.50) | 240.50(207.00-391.75) | 0.733 |
| BNP, ng/L | 178(43-676) | 923.50(118.00-3583.75) | 0.058 |
| Inflammatory biomarker |  |  |  |
| PCT, ng/ml | 0.04(0.01-0.12) | 0.33(0.15-0.51) | < 0.001 |
| CRP, mg/L | 29.38(3.26-66.34) | 63.37(29.31-94.90) | 0.017 |
| ESR, mm/h | 49.67(26.35-78.90) | 41.40(29.15-67.03) | 0.828 |
| SAA, mg/L | 144.15(9.63-452.82) | 610.37(417.31-1990.66) | < 0.001 |
| IL-6, pg/ml | 2.10(1.50-3.24) | 5.40(2.73-9.47) | < 0.001 |
| D-dimer, ug/ml | 0.72(0.33-1.44) | 2.59(0.92-4.29) | 0.001 |
| FDP, ug/ml | 4.32(2.88-5.61) | 5.16(3.77-9.88) | 0.029 |

**Table 4.** Univariate and Multivariate logistic Regression of Factors Associated with Progression to Death

| Variables | Univariate Analysis |  |  | Multivariate Analysis |  |  |
| --- | --- | --- | --- | --- | --- | --- |
|  | OR | 95%CI | P | OR | 95%CI | P |
| CHD | 4.618 | 1.421-15.011 | 0.011 | 2.806 | 0.971-16.795 | 0.039 |
| Lymphocytes | 0.018 | 0.001-0.252 | 0.003 | 0.040 | 0.001-2.306 | 0.045 |
| Monocytes | 0.002 | 0.001-0.127 | 0.004 | 0.061 | 0.001-2.413 | 0.353 |
| NLR | 1.009 | 0.989-1.029 | 0.396 | 1.102 | 0.923-1.051 | 0.637 |
| AST | 1.035 | 1.015-1.055 | 0.001 | 1.026 | 1.000-1.052 | 0.045 |
| PCT | 1.005 | 0.757-1.0335 | 0.972 | 1.123 | 0.594-2.121 | 0.721 |
| SSA | 1.001 | 1-1.002 | 0.005 | 1.021 | 1.001-1.025 | 0.009 |
| IL-6 | 1.007 | 0.971-1.044 | 0.705 | 1.015 | 0.948-1.086 | 0.667 |
| D-dimer | 1.261 | 1.053-1.512 | 0.012 | 1.231 | 1.042-1.456 | 0.015 |
| FDP | 1.068 | 1.010-1.130 | 0.021 | 1.082 | 0.604-1.288 | 0.051 |

## Discussion

With the rapid COVID-19 spread worldwide, the prevention and control of pandemics should come first. Based on the current study, a study of 1,590 patients with COVID-19 pneumonia in China showed that 25.1% of patients had two or more comorbidities, Which means that about a quarter of the infected population suffers from comorbidities(*4*). The rate in our study was a slightly higher (28.8%) than previous studies. The risk of progression to from the mild type to critical type and mortality rate of COVID-19 pneumonia with comorbidities was significantly higher. Furthermore, we observed that the proportion of patients with hypertension is the highest (*8, 9*), and the ratio of patients with diabetes or coronary heart disease is similar. Although there is no statistical difference whether with comorbidities or not between the two groups, Bivariate cox regression logistic regression analysis showed that past medical history of coronary heart disease is one of the risk factors associated with death.

In addition, we observed that the usage rate of low-dose methyl prednisolones, antibiotics, gamma globulin, and albumin in the critical group was significantly higher than that in the mild group. At the same time, we found some interesting characteristics in laboratory indices between the two groups at admission. COVID-19 pneumonia patients with comorbidities have higher percentages of neutrophils, neutrophil ratio, inflammatory biomarkers, and lower percentages of lymphocytes, lymphocyte ratio, and albumin. This also suggests that patients with comorbidities have a high prevalence of progression to severe disease, and are more likely to induce bacterial infections and cause inflammatory storm. Therefore, for such patients, targeted diagnosis and management and early treatment of basic diseases are of great significance for reducing patient mortality and effectively preventing and controlling the epidemic (*10, 11*). The autopsy on patients died of COVID-19 Pneumonia demonstrated that the structure of myocardial cells in patients was intact, and only a small amount of monocyte infiltration in the intercellular substance was observed (*12, 13*). Our study also did not observe a significant increase in the biomarkers that directly cause myocardial damage in critical group. The elevated level of NT-pro BNP in critical group may be related to patients with cardiovascular disease.

Subsequently, we divided patients with comorbidities into the survivor group and the non-survivor group, and compared the general data and laboratory biomarkers between the two groups. Similarly, the proportion of died patients with coronary heart disease was significantly higher than the survivor group, And No significant differences were observed in age and gender between the two groups. Compared with the survivor group, the neutrophil ratio in died patients was significantly increased when they were admitted to the hospital, while the proportion and count of lymphocytes and monocytes were significantly decreased, which indicates that the patient’s immune system is damaged to a certain extent. Some researchers analyzed the peripheral blood of patients by flow cytometry and found that the number of CD4 + and CD8 + T cells in the blood was significantly reduced, but the cell state was over-activated, showing an increase in the number of Th17 and high CD8 ^+^ T cell cytotoxicity(*8, 14, 15*). It is proposed that lymphopenia is a common feature of COVID-19 patients, which may be related to disease severity and mortality (*2, 7, 12*). We conducted a multivariate cox regression analysis on the indices with statistical differences in the univariate analysis. The results also showed that past medical coronary heart disease history and decreased lymphocytes are independent risk factors for death of COVID-19 pneumonia patients with comorbidities.

In order to explore the patients’ inflammatory state, the levels of inflammatory biomarkers were measured. PCT, CRP, SSA, and IL-6 in the patients in the non-survivor group were also significantly increased. Once the activation of inflammatory pathways in patients activated, it was very likely to quickly cause organ failure and eventually threaten life(*16*). Immunological research (*8, 17*) showed that IL-6 is an important pathway to induce inflammatory storms, and patients with COVID-19 virus infection are different from the previous SARS virus, some patients may have early onset and are not severe or even mild symptoms. However, the activation of the later inflammatory storm may accelerate the patient into a state of multiple organ failure, especially in patients with comorbidities. Furthermore, the results of this study showed that the AST, D-dimer, and fibrin degradation products were significantly higher in the non-survivor group than in the survivor group. Previous research have shown that the main causes of liver injury of new coronary pneumonia include immune injury, drug factors, systemic inflammation and ischemia-hypoxia-reperfusion injury (*18*). On the one hand, many patients have already taken one or more antiviral drugs when they were admitted to the hospital. On the other hand, under the condition of shock and hypoxia, there were oxygen deprivation, lipid aggregation, and glycogen depletion in the liver cells, which inhibited cell survival signals, causing liver cell damage and death. In addition, an analysis specifically on the laboratory findings of 82 deaths also showed that, most patients have abnormalities of CRP, lactate dehydrogenase, D-dimer, AST and ALT(*20*). Our multivariate stepwise regression analysis shows that elevated AST, SSA, and D-dimer are risk factors of death in patients with comorbidities. This also reminds us that in the treatment of patients with severe pneumonia, we must not only strengthen the control of inflammatory factor storms, but also pay special attention to the treatment of basic diseases and the protection of organ functions.

Patients with comorbidities in COVID-19 Pneumonia are a special kind of group, and these patients are often in serious condition at admission and have a risk of poor prognosis. Therefore, the evaluation of comorbidities and the establishment of the risk classification of patients with COVID-19 pneumonia upon admission can help to classified management and determine which patients are more likely to have serious adverse outcome in the progression of pneumonia. The results of this study also suggest that we need to pay attention to the treatment of patients’ basic diseases in the future treatment, early intervention in inflammatory storms, and protection of other organs. This study still has many disadvantages. It is only a single-center descriptive study, focusing on the starting point and outcome of patients. In the future, it is necessary to expand the sample size and track the development of disease.

## Data Availability

All data generated or analyzed during this study are included in this article.

## Acknowledgement

None

